# Restrictive Transgender Legislation on Self-Reported Mental Health and Suicidality Results from the Youth Risk Behavior Surveillance Survey, 2010-2019

**DOI:** 10.1101/2025.08.28.25332950

**Authors:** Jasper Kenney, Jennifer Schanzle, Charisse Graham, Fazlur Rahman, Samantha V. Hill

## Abstract

**Objective:** This study examined anti-transgender legislative trends’ impact on high school youth reported violence and suicide-related behaviors.

**Design:** This retrospective analysis collected data from 2010-2019 by the Human Rights Campaign; Legislation Affecting LGBTQ Rights Across the Country; and the Centers for Disease Control and Prevention’s Youth Risk Behavior Surveillance survey.

**Methods:** Violence and suicide outcomes were matched to legislation counts, with frequency distribution summarized by state. Chi-square and Fisher exact tests assessed the association between state responses and number of state passed bills using SAS.

**Results:** One hundred, ninety-two anti-transgender bills were tallied. Twelve bills passed during the study period. As the number of anti-transgender bills passed increased, youth respondents were more likely to report experiencing/engaging in 13 of 17 violence-associated behaviors: (carrying a weapon (p<0.0001), gun (p=0.0009), been threatened/injured by a weapon (p<0.0001), physical fight (p=0.383), bullied at school (p=0.0058) or suicide (serious considerations (p<0.0001), attempt (p=0.0004), treatment after attempt (p=0.0465)). Transgender youth were more likely to report suicide (considerations, planning, attempting, or treatment after an attempt (all with p<0.0001)).

**Conclusion:** Transgender and non-transgender youth reported experiencing more violence and suicidality as more anti-transgender legislation passed. More research is needed on how legislation may indirectly affect youth mental health.

## Introduction

Transgender youth represent a uniquely vulnerable population. The Youth Risk Behavior Surveillance (YRBS) survey, a national high school survey conducted by the Centers for Disease Control and Prevention (CDC), indicates that 1.8% of high school survey respondents are transgender(1). It also reveals transgender youth have disproportionately high rates of experiencing violence, substance use, and suicide attempts (1). Another nationwide study of United States (US) transgender youth who disclosed being transgender at school found 77% reported discrimination, violence, or other mistreatment, and 52% reported a suicide attempt (2).

Furthermore, the growing volume of US state-level legislation aimed at limiting the rights of transgender people may potentiate these concerns. Legislative records maintained by the American Civil Liberties Union (ACLU) reveals 52 such bills were proposed in 2020 compared to 123 in 2021, and 150 in 2022 at the time of this study(3). Similarly, bills pre-empting or revoking nondiscrimination protections for transgender people on the municipal and federal levels, including restrictions on restroom access, participation in athletics, the ability to update identity documents, and access to healthcare, were numerous mid-decade.

Additionally, state policies restricting nondiscrimination protections, health insurance coverage, and changes to identity documents for transgender people have been associated with less utilization of gender-affirming care(4). More recent and direct restrictions on the legality of gender-affirming care for minors threaten access to care that provides significant benefits to transgender youth. Specifically, providers (5–7) and parents (8) have expressed concern about the potential impacts on mental health and suicidality within this population.

While the number of policies being proposed since 2020 has exploded, these policies and their potential impact on youth have been building for the past decade and have yet to be fully explored. An examination of the previous decade’s legislative trends and how their impact on youth, transgender and non-transgender alike, experiencing victimization and mental health outcomes can provide crucial insights into how the impact of these policies affect reported experiences of violence and thoughts about suicide. This study examines proposed and passed anti-trans legislation trends over the decade from 2010-2019 and the potential relationship between reported rates of exposure to violence and suicidality in high school students in the US. We hypothesize there will be an increase in the frequency of all behaviors, and suicidal behaviors specifically, among transgender respondents as the number of anti-transgender bills passed increases.

## Methods

### Design

This retrospective study was deemed non-human subjects research by the University of Alabama at Birmingham secondary to it being a secondary analysis of publicly available data.

### Data Sources

We utilized existing documents logging transgender bills in state legislatures created by advocacy organizations (The Equality from State to State (available for 2004-2013) and State Equality Index(9) (available for 2014-2018) reports by Human Rights Campaign and the Legislation Affecting LGBTQ Rights Across the Country tracker (available for 2018-2022)(10) maintained by the ACLU) to develop a list of anti-transgender bills proposed in each state from 2010-2019(9, 10).

### Data Instruments

Bills were included if they encompassed restrictions specifically based on gender identity or sex assigned at birth. Bills broadly affecting the LGBTQ community as a whole or referencing gender identity in combination with sexual orientation were excluded. The status of each piece of legislation as passed or failed was recorded. Bills introduced in both chambers were tallied individually regardless of whether they were companion bills. Due to variance in state legislative schedules, we tracked the number of bills in a given state legislature each year, regardless of whether they rolled over from a previous year.

The YRBS survey, administered every odd year, is a national survey of high school student behaviors. Each state data set(11–16) as well as the corresponding SAS format and input programs were obtained from the CDC YRBS survey website(11–16). The sample sizes for each survey administration were: 2009-16,410 responses(11), 2013-13,583 responses(12), 2015-15,713 responses(13), 2017-14, 956 responses(14), and 2019-13,677 responses(15). Total number of responses were not available for 2011, but estimated student responses by state ranged from 1,147-13,201 from 47 states(16).

We compared state YRBS survey data to the total number of anti-transgender bills passed every 2 years per state to align with the YRBS survey administration. For example, for the data from state “X” 2017 YRBS survey, the total number of bills for “X” state in 2016 and 2017 were recorded. In 2017, nine states expanded gender classification on their YRBS survey to include transgender youth, with 14 states collecting expanded gender classification in 2019.

### Variables

Dependent variables included responses self-reporting experiencing violence, threats of violence, depression, or suicidal ideation and attempt as reported on the YRBS survey. **Table 1** included YRBS questions. Independent variables included self-reported gender identity (dichotomized to transgender vs not transgender) and number of anti-transgender legislation passed. Gender identity was assessed using the following question as worded by the YRBS survey “Some people describe themselves as transgender when their sex at birth does not match the way they think or feel about their gender. Are you transgender?” Response options included: 1) no, not transgender, 2) yes, transgender, 3) not sure, 4) not understanding the questions. The variable was categorical with individuals who responded yes or unsure being classified as transgender and no being classified as cis-gender. Individuals who did not understand the question were not included in this study. The number of anti-transgender legislation passed was a categorical variable (0, 1, or 2).

**Table 1:**
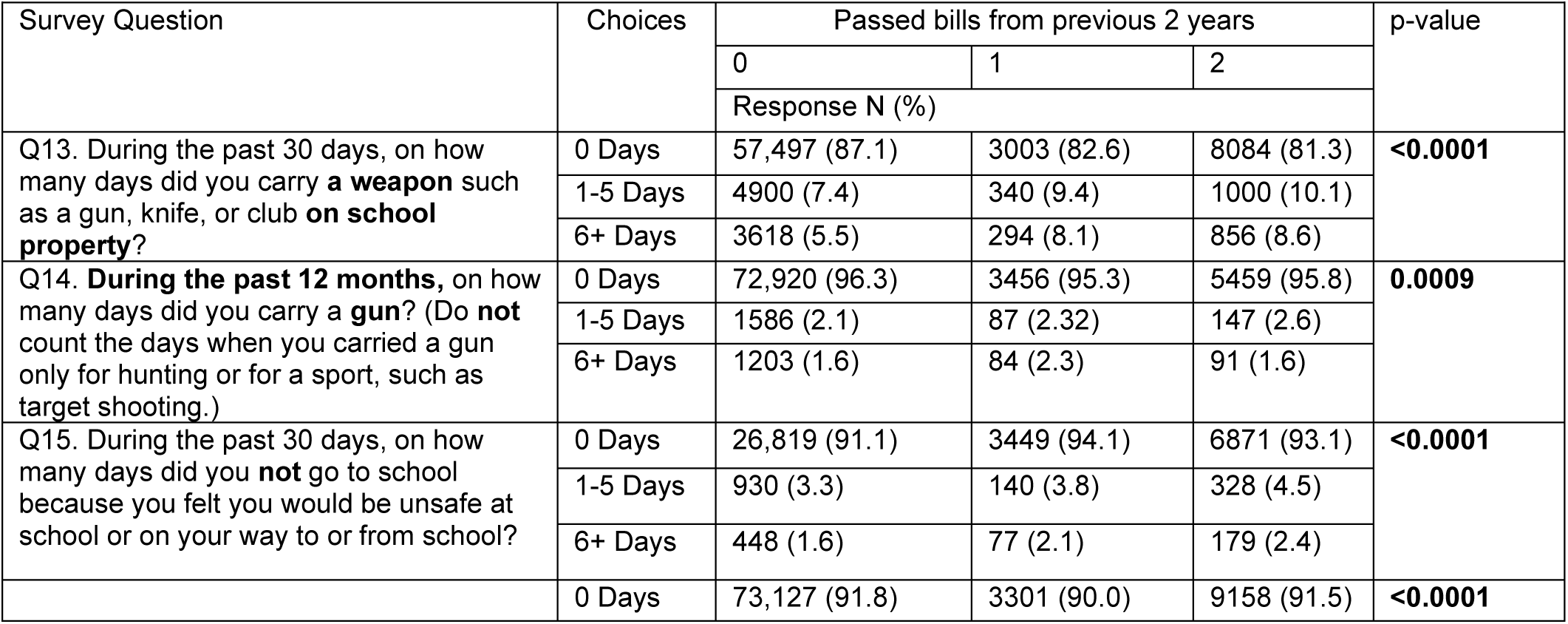
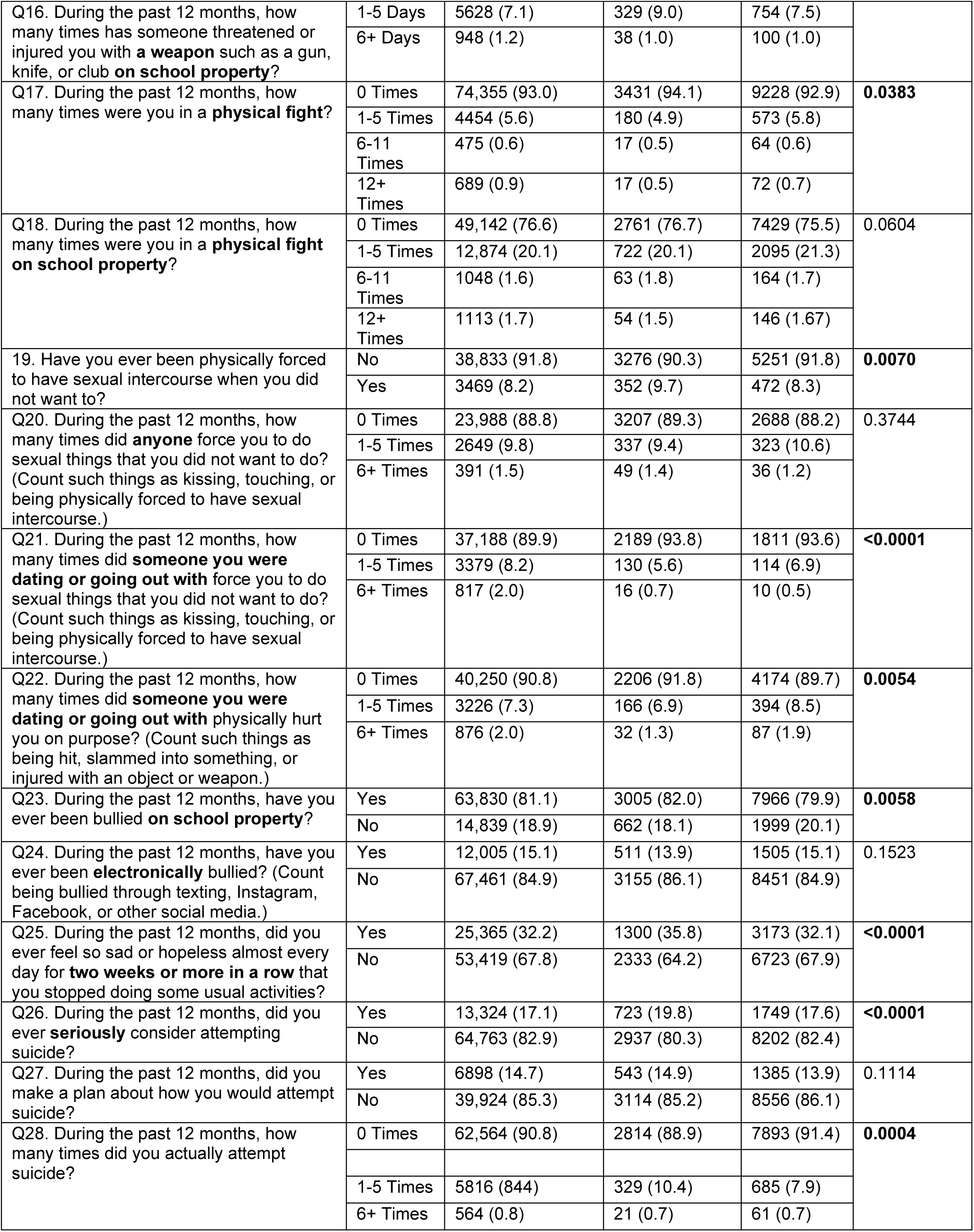
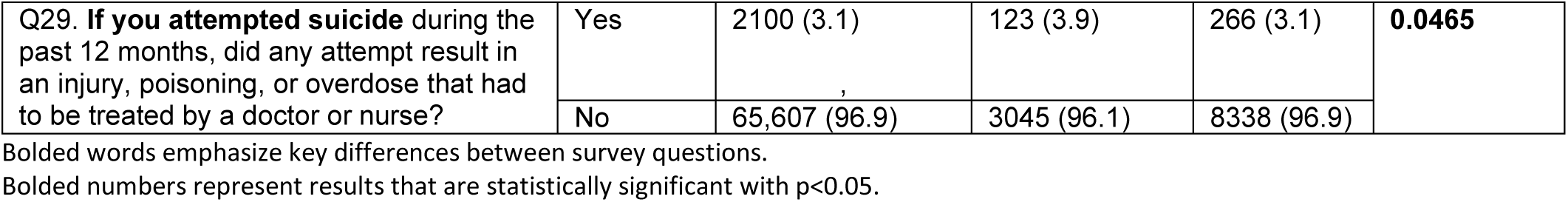
Relationship between Negative Transgender Policies Passed and Youth Reports of Violence, Mental Health, or Suicide on the CDC’s Youth Risk Behavior Surveillance Survey 2010-2019.

### Data Analysis

YRBS survey question responses were matched and summarized by state using frequency distribution (%). State YRBS responses were paired with the number of bills passed by state during the previous two years. Chi-square and Fisher exact tests as appropriate were conducted to assess the association between state responses and number of bills passed by state from the previous two years. Similar analyses were also performed to evaluate associations between selected responses by state and 2017 and 2019 YRBS survey state-level transgender status, specifically. All hypothesis tests were two-tailed with p-value <0.05 used to indicate statistical significance. All analysis used SAS version 9.4 (Cary, NC).

## Findings

192 pieces of anti-transgender bills were proposed between 2010 to 2019. The largest number of bills proposed was in 2016 (55 bills) (**Figure 1**). Twelve bills passed in the decade. **Table 2** illustrates the relationship between the number of bills passed during the 2010-2019 US legislative sessions, and the frequency with which all YRBS survey respondents reported a particular behavior. Respondents were statistically more likely to report experiencing or engaging in 14 behaviors as the number of bills passed increased (bolded in **Table 2**).

**Figure 1.**
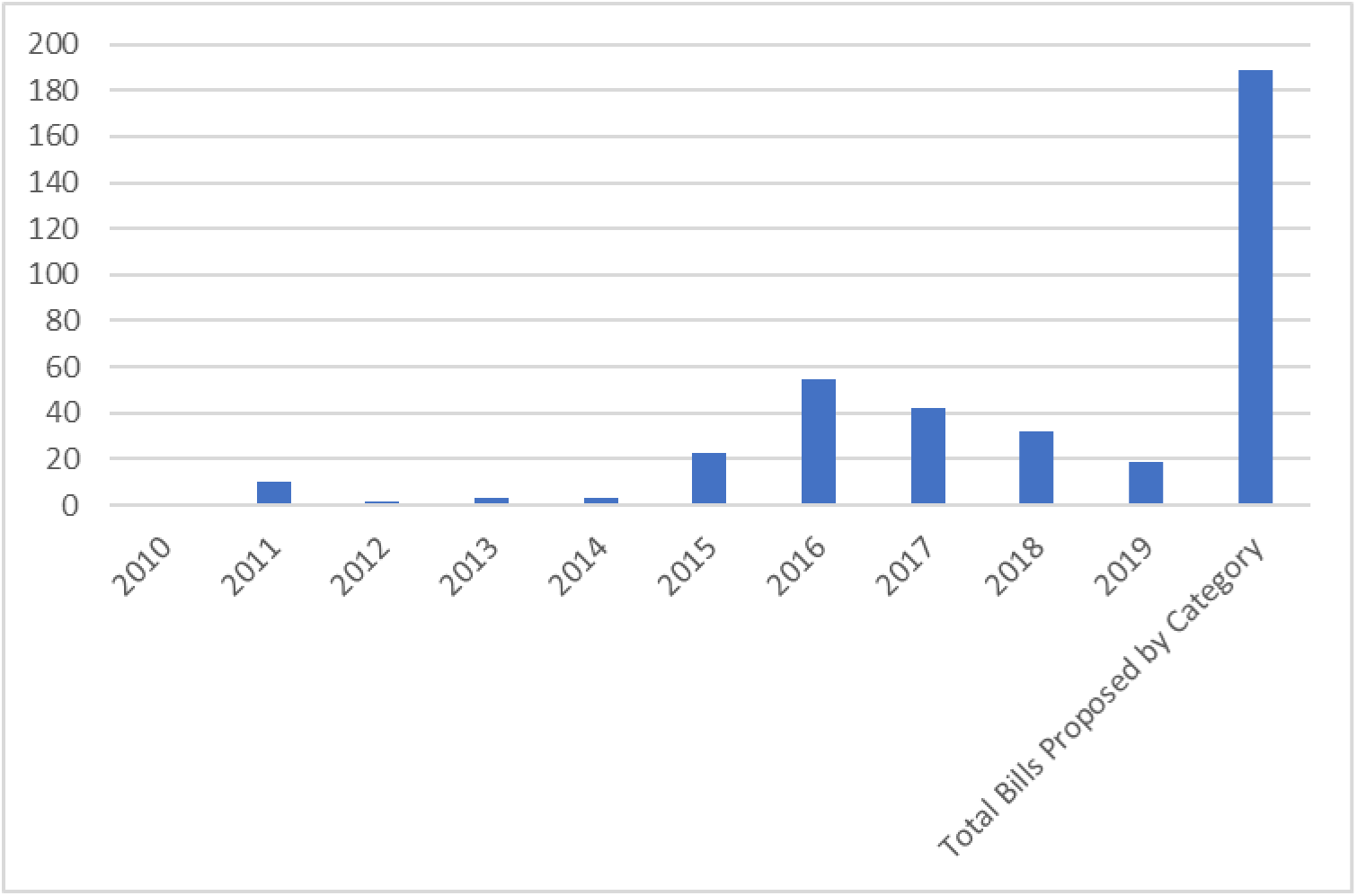
Anti-transgender Legislation Bills Proposed Over Time. This figure displays the number of anti-transgender legislative bills proposed each year during 2010-2019 and the total number of bills proposed during the entire time period.

**Table 2:**
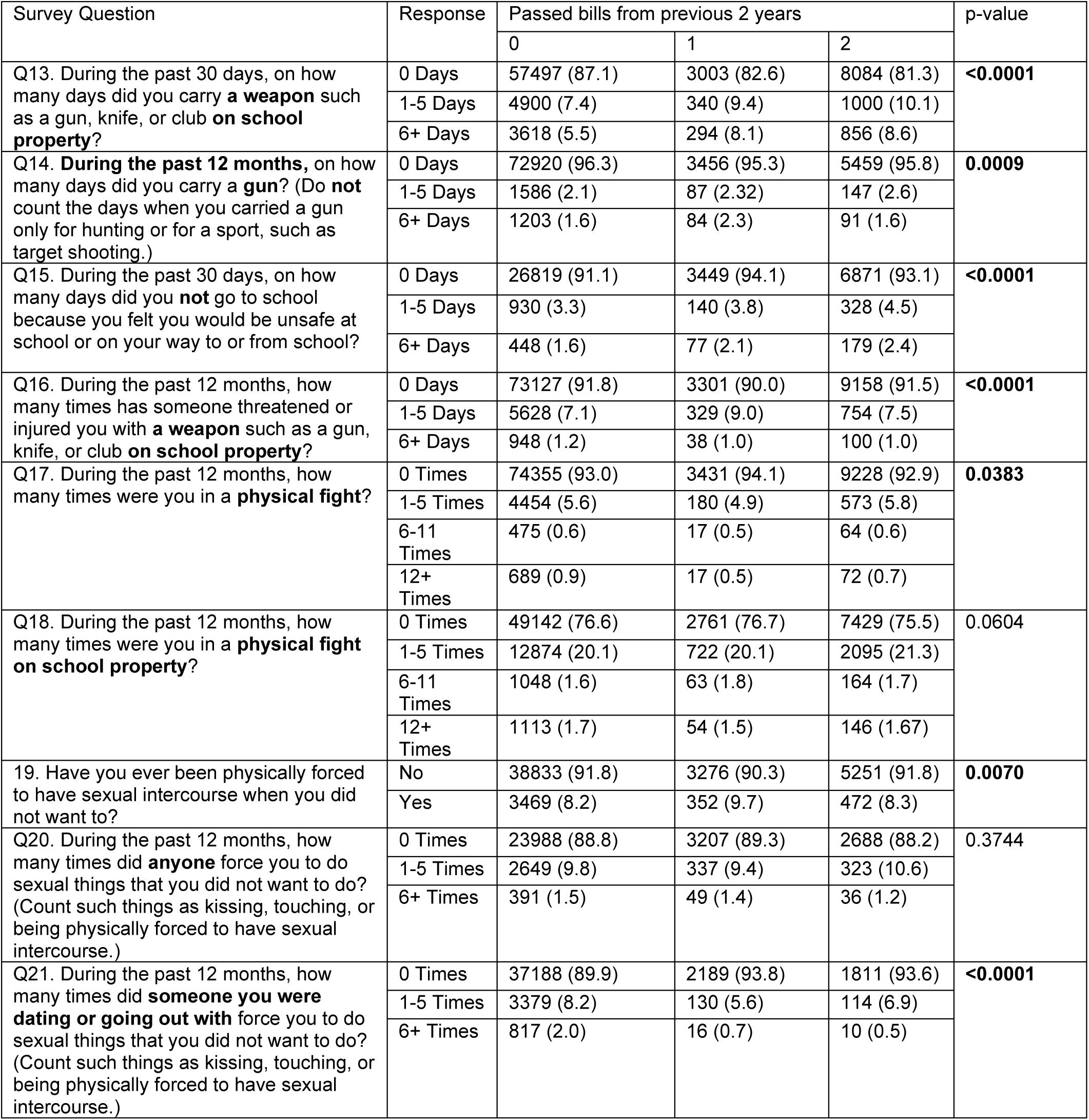
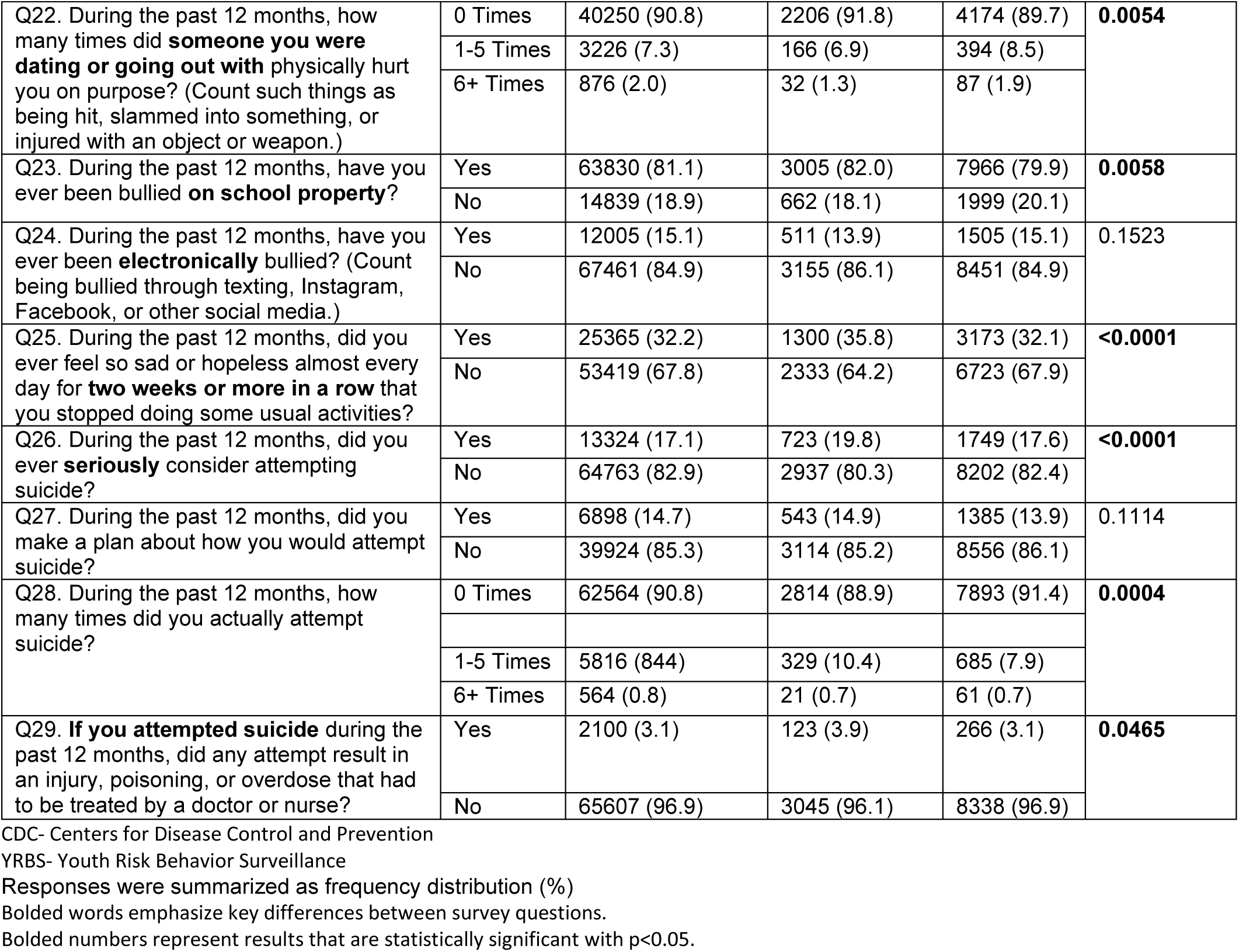
Relationships between Bills Passed and Select Responses to the CDC’s YRBS Survey 2010-2019.

**Table 3** shows the relationship between transgender youth and mental health and suicidal behaviors. Transgender youth were statistically more likely to report experiencing or engaging in the following behaviors as the number of bills passed increased: ever seriously consider attempting suicide (p<0.0001), making a plan about how they would attempt suicide (p<0.0001), actually attempting suicide (p<0.0001), having a suicide attempt result in an injury, poisoning, or overdose that had to be treated by a doctor or nurse (p<0.0001)

**Table 3:** Select Question Responses on Suicidality Stratified by Transgender Status during 2017 and 2019.

| Suicidality Question |  | Year |  |  |  |  |  |  |  |  |
| --- | --- | --- | --- | --- | --- | --- | --- | --- | --- | --- |
|  |  | 2017 |  |  | 2019 |  |  | 2017 and 2019 Combined |  |  |
|  |  | Transgender Yes | Transgender No | p-value | Transgender Yes | Transgender No | p-value | Transgender Yes | Transgender No | p-value |
| Q.26 | Yes | 807 (44.4) | 11528 (16.5) | **<0.0001** | 665 (51.1) | 14833 (17.2) | **<0.0001** | 1472 (47.2) | 26361 (16.9) | **<0.0001** |
| Q. 27 | Yes | 783 (38.5) | 11794 (13.1) | **<0.0001** | 685 (47.2) | 13377 (14.2) | **<0.0001** | 1468 (42.1) | 25171 (13.7) | **<0.0001** |
| Q. 28 | 0 times | 440 (66.1) | 39931 (93.4) | **<0.0001** | 645 (68.8) | 55951 (92.2) | **<0.0001** | 1085 (67.6) | 95882 (92.7) | **<0.0001** |
|  | 1-5 times | 159 (23.9) | 2564 (6.0) |  | 227 (24.2) | 4369 (7.2) |  | 386 (24.1) | 6933 (6.7) |  |
|  | 6+ times | 67 (10.1) | 254 (0.6) |  | 66 (1.0) | 360 (0.6) |  | 133 (8.3) | 614 (0.6) |  |
| Q. 29 | Yes | 37 (18.6) | 316 (2.6) | **<0.0001** | 74 (17.2) | 753 (2.5) | **<0.0001** | 111 (17.6) | 1069 (2.5) | **<0.0001** |

Bolded words emphasize key differences between survey questions. Bolded numbers represent results that are statistically significant with $p < 0.05$ .

## Discussion

This study explores the historical relationship between anti-transgender legislation and youth (transgender and non-transgender) violence and suicide behaviors using a decade of legislative data. Findings revealed the amount of anti-transgender bills roughly increased until their peak in 2016 with the frequency of behaviors such as not attending school, being physically or sexually assaulted, feeling sad and considering suicide increasing as the number of bills passed increased. Surprisingly, increased considerations for suicide as the number of antitransgender bills passed was noted among all youth, regardless of gender identity.

This study adds to literature that supports a potential link between policies broadly focused on restricting the rights of the LGBTQ community and increased violence, victimization experiences, and negative mental health outcomes among youth. For instance, a 2012 Massachusetts state law establishing nondiscrimination protections excluding discrimination in public accommodations based on gender identity has been associated with both high rates of discrimination and adverse emotional and physical health effects experienced by transgender people(17). A 2023 survey found one-third of LGBTQ youth reported poor mental health as a result of anti-LGBTQ policies and legislation(18). In 2024, the Trevor project found 47% and 52% of surveyed transgender young women and men respectively considered suicide in the past year with rates double that of their cis-gender peers, with qualitative data revealing youth are relating these feelings to the current legislative climate.

Contrastingly, the legality of same-sex marriage has also been associated with reduced youth suicidality both on the state level within the US (19) and internationally (20), with affirming legislative climates and higher community LGBTQ-supportiveness having been associated with lower rates of suicidal ideation, suicide attempts, and self-harm among sexual minority adolescent girls (21).

Most surprisingly, reports of sadness, considerations of suicide, and suicide attempts were increased among both transgender and non-transgender youth alike as the number of anti-transgender bills passed increased. Data is lacking regarding how anti-transgender legislation specifically impacts non-transgender youth. Most available data focus on how school policies impact LGBTQ youth (22). However, one California study (23) did reveal that LGBTQ inclusive policies resulted in all students feeling safer and experiencing less victimization. More prospective studies examining how anti-transgender policy impacts both transgender and non-transgender youth are needed.

The direct impacts of anti-transgender policies and the general social and political climate to which such policies contribute are important considerations for clinicians, especially regarding the politicization of transgender youth. This includes increasing concerns around bans on transgender youth participation in sports and potential consequences to the well-being of transgender youth given these pieces of legislation(24). Additionally, even though evidence-based clinical guidelines indicate that treatment of gender dysphoria through gender-affirming care has benefits including improvements in anxiety, depression, and suicidality, state-level restrictive policies have dramatically increased in this area(4, 25). Thus, it is not surprising that our study, like others, found that policies restricting the rights of transgender people would have similar effects across the spectrum of the LGBTQ community.

### Limitations

While this study provides additional insights into the potential relationship between anti-trans legislation and youth behavior, there are some limitations. There was not a centralized, continuous record of legislation and data was aggregated from various sources (Human Rights Campaign and ACLU), which introduces opportunities for subjectivity or error. Additionally, some bills spanned multiple categories and may have complicated the classification process.

Variation in the way state legislatures function with respect to length of sessions and whether bills from one chamber can be directly approved by another chamber without a distinct companion bill make it challenging to account for national and state-specific time effects and confounding variables. Furthermore, bills do not need to explicitly mention transgender people to impact them deeply, as transgender people are more likely than their peers to be living in poverty or to be incarcerated(2). Any legislation impacting state resources for low-income residents or the state criminal justice system might disproportionately impact transgender people as well. This study was not able to account for factors such as these in compiling legislation.

Use of YRBS survey data to represent youth nationally posed additional limitations. Not all states participate in the YRBS survey, and some do not have data available for certain years due to an insufficient number of responses. Further complicating this is that questions about gender identity and transgender status only became part of the standard question set beginning in 2017. Additionally, states can and sometimes do choose to omit these items from their statewide questionnaires. Similarly, the methodology of the study introduces difficulty with drawing direct conclusions given the mapping of behaviors with respect to legislation as opposed to surveying subjects about their experiences following introduction and passage of bills.

## Conclusion

This study provides a historical look into how anti-transgender legislation impacts mental health and experience of victimization among both transgender and non-transgender youth alike. As the number of anti-transgender bills proposed and passed has soared since 2020, understanding the potential impact of these bills have on all youth is crucial. More studies are needed to understand the temporality between increases in experiencing victimization and considering suicidality as sequalae of anti-transgender legislation.

## Supporting information

Strobe Checklist

## Data Availability

All data produced in the present study are available upon reasonable request to the authors. Requests for publicly accessible data can be sent to the organizations listed above.

## Abbreviations

YRBS: Youth Risk Behavior Surveillance
CDC: Centers for Disease Control and Prevention
LGBTQ: Lesbian, Gay, Bisexual, Transgender, and Queer
U.S.: United States
ACLU: American Civil Liberties Union

## Declarations

### Ethics Approval

Ethical approval for this study was obtained from the University of Alabama at Birmingham Institutional Review Board (approval number: IRB-981112002)

### Consent for publication

Not applicable

### Authorship confirmation

This study was conceptualized by JK and SH. The study procedures were developed by JK, SH, and FR. The analysis, development of the manuscript, and final approvals of the manuscript were from JK, SH, FR, CG, and JS.

## Acknowledgements

We would like to thank the Centers for Disease Control and Prevention for providing us with granular data from their Youth Risk Behavior Surveillance Survey.

## References

1. Johns MM, Lowry R, Andrzejewski J, Barrios LC, Demissie Z, McManus T, et al. Transgender identity and experiences of violence victimization, substance use, suicide risk, and sexual risk behaviors among high school students - 19 states and large urban school districts, 2017. MMWR Morb Mortal Wkly Rep. 2019;68(3):67–71.

2. James SE, Herman, J. L., Rankin, S., et al. The Report of the 2015 U.S. Transgender Survey. Washingtong, DC: National Center for Transgender Equality.; 2016.

3. Kimball AA, Zhu W, Leonard J, Wei W, Ravichandran I, Tanner MR, et al. HIV preexposure prophylaxis provision among adolescents: 2018 to 2021. Am Acad Pediatr. 2023;152(5).

4. Goldenberg T, S LR, G WH, K EG, Stephenson R. State-level transgender-specific policies, race/ethnicity, and use of medical gender affirmation services among transgender and other gender-diverse people in the United States. Milbank Q. 2020;98(3):802–46.

5. Hughes LD, Kidd KM, Gamarel KE, Operario D, Dowshen N. “These laws will be devastating”: provider perspectives on legislation banning gender-affirming care for transgender adolescents. J Adolesc Health. 2021;69(6):976–82.

6. Kremen J, Williams C, Barrera EP, Harris RM, McGregor K, Millington K, et al. Addressing legislation that restricts access to care for transgender youth. Am Acad Pediatr. 2021;147(5):1–4.

7. Barbee H, Deal C, Gonzales G. Anti-transgender legislation-a public health concern for transgender youth. JAMA pediatrics. 2022;176(2):125–6.

8. Kidd KM, Sequeira GM, Paglisotti T, Katz-Wise SL, Kazmerski TM, Hillier A, et al. “This could mean death for my child”: parent perspectives on laws banning gender-affirming care for transgender adolescents. J Adolesc Health. 2021;68(6):1082–8.

9. Equality from State to State & State Equality Index Archives Washington, DC: Human Rights Campaign; [Available from: https://www.hrc.org/resources/equality-from-state-to-state.

10. Legislation Affecting LGBTQ Rights Across the Country New York, NY: American Civil Liberties Union; 2024 [Available from: https://www.aclu.org/legislation-affecting-lgbtq-rights-across-country

11. 2009 High School Youth Risk Behavior Survey Data (SAS Dataset) Atlanta, GA: Centers for Disease Control and Prevention (CDC).; 2009 [updated April 2021. Available from: http://yrbs-explorer.services.cdc.gov/.

12. 2013 High School Youth Risk Behavior Survey Data (SAS Dataset) Atlanta, GA: Centers for Disease Control and Prevention (CDC).; 2013 [updated April 2021. Available from: http://yrbs-explorer.services.cdc.gov/.

13. 2015 High School Youth Risk Behavior Survey Data (SAS Dataset) Atlanta, GA: Centers for Disease Control and Prevention (CDC). 2015 [updated April 2021. Available from: http://yrbs-explorer.services.cdc.gov/.

14. 2017 High School Youth Risk Behavior Survey Data (SAS Dataset) Atlanta, GA: Centers for Disease Control and Prevention (CDC).; 2017 [updated April 2021. Available from: http://yrbs-explorer.services.cdc.gov/.

15. 2019 High School Youth Risk Behavior Survey Data (SAS Dataset) Atlanta, GA: Centers for Disease Control and Prevention (CDC). 2019 [updated April 2021. Available from: http://yrbs-explorer.services.cdc.gov/.

16. 2011 High School Youht Risk Behavior Survey (SAS Dataset) Atlanta, GA: Centers for Disease Control and Prevention (CDC); 2011 [updated April 2021. Available from: http://yrbs-explorer.services.cdc.gov/.

17. Reisner SL, Hughto JM, Dunham EE, Heflin KJ, Begenyi JB, Coffey-Esquivel J, et al. Legal protections in public accommodations settings: A critical public health issue for transgender and gender-nonconforming people. Milbank Q. 2015;93(3):484–515.

18. 2023 U.S. National Survey on the Mental Health of LGBTQ Young People West Hollywood, CA: The Trevor Project 2023 [Available from: https://www.thetrevorproject.org/survey-2023/.

19. Raifman J, Moscoe E, Austin SB, McConnell M. Difference-in-differences analysis of the association between state same-sex marriage policies and adolescent suicide attempts. JAMA pediatrics. 2017;171(4):350–6.

20. Kennedy A, Genc M, Owen PD. The association between same-sex marriage legalization and youth deaths by Suicide: A multimethod counterfactual analysis. J Adolesc Health. 2021;68(6):1176–82.

21. Saewyc EM, Li G, Gower AL, Watson RJ, Erickson D, Corliss HL, et al. The link between LGBTQ-supportive communities, progressive political climate, and suicidality among sexual minority adolescents in Canada. Prev Med. 2020;139:106191.

22. Arcelo JM, Delim, Maria Claudia, Eribal, Drixel Vherniz, et al. Evaluating LGBTQIA acceptance: An exploratory study smong LGBTQ and non-LGBTQ students in selected sectarian schools and universities. Int Multidiscip Res J. 2023;Volume 6(Issue 6):pp. 222–30.

23. McGovern AE. When schools refuse to say gay: The constitutionality of anti-LGBTQ no-promo-homo public school policies in the United States. JL & Pub Policy. 2012;Vol. 22( Iss. 2):1–26.

24. Hughes LD, Dowshen N, Kidd KM, Operario D, Renjilian C, Gamarel KE. Pediatric provider perspectives on laws and policies impacting sports participation for transgender youth. LGBT health. 2022;9(4):247–53.

25. Kraschel KL, Chen A, Turban JL, Cohen IG. Legislation restricting gender-affirming care for transgender youth: Politics eclipse healthcare. Cell reports Medicine. 2022;3(8):100719.

